# Depression, immune–metabolic alterations and mortality in haemodialysis: a prospective cohort study

**DOI:** 10.64898/2026.07.29.26359196

**Authors:** Claire Duperrex, Gemma Smith, Christophe Clesse, Olivier Duperrex, Simone Rahman, Georgina M. Hosang, Anastasia Prokopi, Brian Gracey, Fiona Loud, Simon Kirwin, David Randall, Karl Marlowe, Steve Cole, Kamaldeep Bhui, Magdi Yaqoob, Livia A Carvalho

## Abstract

**Introduction:** Depression is common among people receiving haemodialysis and is associated with adverse outcomes, but the biological pathways underlying this relationship remain uncertain. We examined associations between depressive symptoms, mortality, immune–metabolic markers and stress-related transcriptional profiles, including potential sex differences in routine biomarkers.

**Methods:** We studied 297 adults receiving maintenance haemodialysis across North and East London and Essex. Moderate/severe depressive symptoms were defined as a 17-item Hamilton Depression Rating Scale score >18. Cox regression examined all-cause mortality from enrolment until death or administrative censoring on 24 March 2026. Linear regression assessed associations between depressive-symptom severity and routinely measured immune–metabolic markers, overall and by sex. In a laboratory subset, inflammatory proteins and transcriptional profiles were examined overall only.

**Results:** Moderate/severe depressive symptoms were associated with higher mortality in the adjusted model, although the association was attenuated after full adjustment (hazard ratio, 1.49; 95% confidence interval, 1.00–2.23; P=0.052). Higher white-cell count was associated with greater depressive-symptom severity before, but not after, adjustment for body mass index. Diastolic blood pressure was the only routine marker showing evidence that its association with depressive symptoms differed by sex (interaction P=0.022). Exploratory analyses did not provide convincing evidence that the measured biomarkers accounted for the depression–mortality association. In the laboratory subset, no inflammatory protein or transcriptional measure was significantly associated with depressive symptoms in fully adjusted analyses.

**Conclusion:** Depression may identify a clinically relevant risk state among people receiving haemodialysis. The exploratory biological findings require validation in larger, prospectively sampled cohorts.

## Introduction

Chronic kidney disease (CKD), and particularly end-stage kidney disease requiring haemodialysis, is a clinically severe long-term condition with a substantial psychological and social burden. CKD also frequently coincides with health inequalities: diabetes mellitus, hypertension and obesity contribute to risk, but do not fully explain the excess burden of kidney disease observed in socioeconomically disadvantaged and ethnical minority populations ^1–5^. Depression is also highly prevalent and might be a marker of cumulative biological and psychosocial vulnerability ^6^. Clinically significant depressive symptoms affect approximately one quarter of people with CKD and is highly comorbid to CKD risk factors (e.g. diabetes, hypertension and cardiovascular disease) ^7–10^.

The high prevalence of depression in CKD is clinically important because depressive symptoms are associated with mortality, and other adverse clinical outcomes ^6^. Standard antidepressant treatment has shown only modest benefits ^11^ in haemodialysis populations, suggesting that depression in CKD may be partly driven by biological and environmental mechanisms that are not adequately targeted. A more integrated biopsychosocial framework is therefore needed, particularly in socially diverse renal populations in which chronic illness, deprivation, stigma, social isolation and repeated exposure to threatening life events may converge.

One plausible mechanism linking adverse social environments, depression and CKD progression is stress-related immune dysregulation. Chronic psychosocial stress disrupts the neuroendocrine, central nervous and immune systems, including cardiovascular, hypothalamic-pituitary-adrenal axis signalling and inflammatory responses ^12^, and may contribute to increased allostatic load over time ^13,14^. In depression, convergent evidence implicates low-grade inflammation and altered cell-mediated immunity, with elevations in inflammatory markers such as C-reactive protein (CRP) reported across multiple studies ^15–21^. Prior studies in CKD have also reported higher immune-metabolic burden among patients with depressive symptoms compared with those without depression ^22^. This overlap raises the possibility that, in CKD, depressive symptoms and adverse clinical outcomes may share stress-response biology. However, it remains unclear whether routinely available clinical biomarkers, such as CRP, white cell count (WCC), lipids and blood pressure, adequately capture the immune-metabolic phenotype associated with depression in haemodialysis populations, and whether this phenotype differs by sex. Inflammatory profile associated with depression may reduce heterogeneity and point towards a subgroup that may be more vulnerable to morbidity and mortality and could potentially benefit from targeted approaches.

Beyond circulating biomarkers, the conserved transcriptional response to adversity (CTRA) describes a stress-related gene-expression profile characterised by upregulation of pro-inflammatory genes and downregulation of genes involved in type-I interferon (IFN) antiviral responses and antibody synthesis ^23–25^ associated with social adversity. Although potentially adaptive under acute threat, persistent activation of this programme may promote chronic inflammatory activity and long-term disease vulnerability. In CKD, where baseline inflammatory burden is already elevated, CTRA-related transcriptional changes may help identify a subgroup of patients in whom depressive symptoms reflect stress-related immune activation rather than psychological distress alone. We thus tested the specific hypothesis that depressed people receiving haemodialysis show disturbances in markers of the CTRA, an RNA profile characterised by up-regulated inflammation and down-regulated IFN/antiviral activity.

Sex may further modify these associations. Women have a higher lifetime prevalence of depression and show distinct immune and hormonal regulation compared with men. Women are disproportionally affected by depression and may be more vulnerable to the depressogenic effects of inflammation than men ^26^. Experimental and epidemiological evidence suggests sex differences in immune-metabolic responses to stress and in the temporal relationship between such disturbances and depressive symptoms ^27–30^. In CKD, sex-specific differences in immune activation, hormonal milieu, autoimmune risk and social adversity exposure may therefore influence the association between immune impairment and depression. Testing sex-stratified associations is consequently important for determining whether a single biomarker model is appropriate across the haemodialysis population.

The present study examined the relationship between depressive symptoms, mortality, stress-response biomarkers and immune/antiviral-related transcriptional signatures in a multi-ethnic cohort of people receiving haemodialysis in North and East London and Essex. We investigated whether depression was associated with 4-years mortality. We also tested whether routinely collected immune-metabolic biomarkers were associated with moderate/severe depression, overall and stratified by sex. We hypothesised that:

1. moderate/severe depression in haemodialysis is associated with premature mortality;
2. routinely collected stress-related markers partly explain this association;
3. deeper inflammatory/transcriptional profiling captures additional stress-related disturbances.

## Methods

### Participants

Of 300 haemodialysis patients recruited across Barts Health NHS Trust between October 2021 and September 2022, serving socioeconomically disadvantaged and ethnical minority community, 297 had linked NHS clinical, biochemistry and mortality data. The renal service at Barts Health NHS Trust runs haemodialysis units at hospitals in North and East London and Essex in the UK (including the Royal London Hospital, Newham General Hospital, King George’s Hospital, Queens Hospital and Whipps Cross Hospital). The service provides care for people of diverse ethnic backgrounds in one of the most deprived parts of the UK. We interviewed the 300 consenting participants and reviewed their clinical records. Participation was voluntary, and patients could withdraw at any stage. We had access to linkage hospital data where information on clinical (type of CKD, diabetes, hypertension, mortality status), sociodemographic (age, gender, weight, height, ethnicity) and stress response biomarkers (CRP, lipid profile, glycated haemoglobin (HbA1c), WCC, systolic blood pressure (sBP), diastolic blood pressure (dBP)). We provided study information to potential participants during haemodialysis and sought written informed consent. All procedures involving human subjects/patients were approved by Health Care Research Wales (reference 19/LO/1272; IRAS project ID 256696). Queen Mary University of London was the sponsor.

### Measures of Moderate/Severe Depression

We used a modified version of the 17-item version of the HAMD-17 ^6,31^ to calculate an overall score. Moderate/severe depression (“Depressed” in tables and plots) was classified as HAMD-17 > 18. An experienced clinical psychologist conducted the interviews (CC). We also assessed anxiety symptoms for common validated screening instruments that are used in routine clinical practice and research (General Anxiety Disorder-7 (GAD), List of Threatening Experiences (LTE)). A smaller subset of 42 individuals with and without moderate/severe depression were invited for a more in-depth lab analysis and provided a follow-up HAMD-17.

### All-cause Mortality

We obtained the survival status of all included participants censoring on the date 2026-03-24. We calculated the follow-up time from the date of study consent (index date) to the date of death or the censoring date, whichever occurred first. We right-censored participants alive at the end of the study period to write this paper.

### Data Cleaning and Outlier Detection

Prior to modelling, we screened 11 continuous variables for data errors using three criteria: clinical plausibility limits, interquartile range (1.5×IQR), and z-scores (|z| > 3). No values were deemed biologically impossible. We identified 36 strong and 60 weak statistical outliers across eight and nine variables, respectively which were retained for analysis to inform the imputation model. All continuous biomarker values were natural-log transformed before analysis to address right skew. Age and body-mass index (BMI) were mean-centred for the models (cAge, cBMI).

### Missing Data Handling

We handled missing data using multiple imputation by chained equations (MICE) with predictive mean matching for numeric variables (m = 20 imputations, 5 iterations) ^32^. Imputation was performed for nine variables: cBMI, CRP, total cholesterol, high-density lipoprotein cholesterol (HDL), low-density lipoprotein cholesterol (LDL), triglycerides, WCC, sBP, and dBP. We confirmed convergence via visual inspection of trace plots. We derived pooled estimates using Rubin’s rules. Complete-case analysis (n = 215/297, 72%) served as a sensitivity check (see Supplementary Materials).

### Descriptive and Primary Analyses

We summarised demographic characteristics, social variables (including life events), and clinical conditions overall and stratified by sex and by BMI categories. Depression was explored as a continuous variable (HAMD-17 score) and as a binary outcome as described above.

Associations between nine exposure variables (CRP, total cholesterol, HDL, LDL, triglycerides, WCC, HbA1c, sBP, dBP) and the continuous HAMD-17 score were examined using linear regression (n = 297). We fitted three nested models for each exposure: (1) unadjusted; (2) adjusted for gender, cAge, ethnicity, CKD, and smoking; and (3) fully adjusted, additionally controlling for cBMI.

### Survival Analyses

All-cause mortality was analysed using Kaplan–Meier estimation and Cox proportional hazards regression, pooled across the 20 imputed datasets. We constructed three nested models (unadjusted, adjusted, fully adjusted) and verified the proportional hazards assumption using Schoenfeld residuals. In all survival models, survival status was treated as observed rather than imputed: the 12 participants with indeterminate vital status were excluded, so hazard ratios rest solely on observed events (n=285). Multiple imputation was applied to covariates only.

For any stress-related biomarker that was significantly associated with depression, we explored whether the depression and mortality association was partly explained by this biomarker in an exploratory mediation analysis. Because vital status was included in the imputation model, the primary mediation decomposition was estimated on the full imputed sample (n = 297), which includes model-based vital status for the 12 participants with indeterminate outcome. This is deliberately less conservative than the survival analysis, which used observed survival status only; mediation is treated here as an exploratory secondary analysis, and the complete-case decomposition (observed outcomes only) is reported as a sensitivity check. This was done using the CMAverse package ^33^ within a counterfactual framework, we estimated both 2-way (natural direct and indirect effects) and 4-way decompositions ^34^. Sensitivity to unmeasured confounding was assessed using E- values ^35^. These analyses were considered exploratory rather than formal causal mediation models, because the observational design and timing of biomarker assessment limit inference about temporal ordering.

### Exploratory Transcriptomic Analysis

In an exploratory analysis, we examined associations between CTRA related gene expression using linear regression with the subset of participants with available transcriptomic data (n total = 42, moderate/severe depression (n=11), no/mild depression (n=31)), applying the same adjustment levels as the primary analysis. Details for the methods are provided in the Supplementary material.

### Data Management and Statistical Analysis

Data were extracted from clinical records, linked to patient interviews, anonymised and stored in Excel. We performed all analyses in R 4.6.0 (R Core Team, 2026) and RStudio (Posit team, 2026), using several key packages: mice for multiple imputation, CMAverse for causal mediation analysis, survival and survminer for time-to-event analyses, easystats for model comparison (see Suppl Table S0). Artificial intelligence tools (Claude, Anthropic; Euria, Infomaniak) were used to support with code writing and were disclosed in accordance with ICMJE recommendations.

## Results

### Prevalence at recruitment

We first examined the prevalence of depressive symptoms in 297 individuals with CKD receiving haemodialysis. Table 1 presents the social, demographic, and clinical characteristics of the cohort stratified by depression status post-imputation. Using the predefined HAMD >18, 83 (27.9%, 95% CI 23.1 to 33.3) participants were classified as moderate/severe depression (“Depressed” in tables and plots) (Mean = 23.54, SD = 4.06) and 214 as no/mild depression (“Not depressed” in tables and plots) (Mean = 9.71, SD = 4.57, p < 0.001). GAD and LTE were both significantly higher in those with moderate/severe depression (p < 0.001 and p = 0.006, respectively). The proportion of individuals deceased by the censoring date did not differ between moderately/severely (45.8%, 95% CI 34.9-57.0) vs not/mildly (54.2%, 95% CI 43.0-65.1) depressed participants (p = 0.102). Moderate/severe depression was more frequent in females than in males, with females representing 53.0% (95% CI 41.8-63.9) of this group compared with 36.9% (95% CI 30.5-43.8) of the not/mildly depressed group (p = 0.012). There were no significant differences by depression on survival status, age, ethnicity, CKD type, smoking, biomarkers or BMI. Overall, these descriptive findings indicate that severe depressive symptoms were common in this haemodialysis cohort and were associated with female sex.

**Table 1:** Social, demographic and clinical characteristics of individuals with CKD by depression status, post-imputation^1^.

| Characteristic<sup>2,3</sup> | Overall<br>N = 297 | Not<br>depressed<br>N = 214 | Depressed<br>N = 83 | p-value |
| --- | --- | --- | --- | --- |
| HAMD-17 total score<sup>4</sup> | 13.58 (7.63) | 9.71 (4.57) | 23.54 (4.06) | <0.001<sup>5</sup> |
| GAD total score<sup>6</sup> | 3.79 (4.82) | 2.21 (3.54) | 7.87 (5.30) | <0.001<sup>5</sup> |
| LTE total score<sup>7</sup> | 1.69 (1.20) | 1.57 (1.13) | 1.98 (1.32) | 0.006<sup>5</sup> |
| Survival status |  |  |  | 0.102<sup>8</sup> |
| Alive | 183 (61.6%) | 138 (64.5%)<br>[57.6-70.8] | 45 (54.2%)<br>[43.0-65.1] |  |
| Deceased | 114 (38.4%) | 76 (35.5%)<br>[29.2-42.4] | 38 (45.8%)<br>[34.9-57.0] |  |
| Gender |  |  |  | 0.012<sup>8</sup> |
| Male | 174 (58.6%) | 135 (63.1%)<br>[56.2-69.5] | 39 (47.0%)<br>[36.1-58.2] |  |
| Female | 123 (41.4%) | 79 (36.9%)<br>[30.5-43.8] | 44 (53.0%)<br>[41.8-63.9] |  |
| Age | 60.39 (15.18) | 61.21 (14.90) | 58.30 (15.78) | 0.186<sup>5</sup> |
| Ethnicity |  |  |  | 0.564<sup>8</sup> |
| White | 89 (30.0%) | 65 (30.4%)<br>[24.4-37.1] | 24 (28.9%)<br>[19.7-40.1] |  |
| Asian | 108 (36.4%) | 74 (34.6%)<br>[28.3-41.4] | 34 (41.0%)<br>[30.5-52.3] |  |
| Black | 100 (33.7%) | 75 (35.0%)<br>[28.7-41.9] | 25 (30.1%)<br>[20.8-41.3] |  |
| Chronic kidney disease type |  |  |  | 0.904<sup>8</sup> |
| Uncertain | 103 (34.7%) | 75 (35.0%)<br>[28.7-41.9] | 28 (33.7%)<br>[24.0-45.0] |  |
| Autoimmune | 45 (15.2%) | 33 (15.4%)<br>[11.0-21.1] | 12 (14.5%)<br>[8.01-24.3] |  |
| Diabetic (Type II) | 64 (21.5%) | 43 (20.1%)<br>[15.1-26.2] | 21 (25.3%)<br>[16.7-36.2] |  |
| Hypertensive | 67 (22.6%) | 50 (23.4%)<br>[18.0-29.7] | 17 (20.5%)<br>[12.7-31.0] |  |
| Polycystic | 18 (6.1%) | 13 (6.1%)<br>[3.41-10.4] | 5 (6.0%)<br>[2.24-14.1] |  |
| Smoking (ever) | 100 (33.7%) | 66 (30.8%)<br>[24.8-37.6] | 34 (41.0%)<br>[30.5-52.3] | 0.098<sup>8</sup> |
| CRP (mg/L)<sup>9</sup> | 11.84 (18.01) | 10.94 (16.88) | 14.17 (20.56) | 0.116<sup>5</sup> |
| Total cholesterol (mmol/L) | 3.89 (1.15) | 3.85 (1.13) | 4.00 (1.18) | 0.304<sup>5</sup> |
| HDL cholesterol (mmol/L)<sup>10</sup> | 1.21 (0.45) | 1.22 (0.46) | 1.21 (0.40) | 0.789<sup>5</sup> |
| LDL cholesterol (mmol/L)<sup>11</sup> | 2.01 (0.92) | 1.99 (0.93) | 2.06 (0.91) | 0.369<sup>5</sup> |
| Triglycerides (mmol/L) | 1.84 (1.61) | 1.77 (1.39) | 2.03 (2.08) | 0.673<sup>5</sup> |
| White cell count (×10<sup>9</sup>/L) | 7.09 (2.80) | 6.87 (2.58) | 7.68 (3.23) | 0.091<sup>5</sup> |
| HbA1c (mmol/mol)<sup>12</sup> | 43.68 (16.31) | 42.96 (16.19) | 45.54 (16.57) | 0.118<sup>5</sup> |
| Systolic blood pressure (mmHg) | 148.99<br>(23.78) | 147.47 (23.31) | 152.93<br>(24.68) | 0.086<sup>5</sup> |
| Diastolic blood pressure (mmHg) | 76.75 (14.27) | 75.87 (13.30) | 79.02 (16.40) | 0.317<sup>5</sup> |
| BMI (kg/m<sup>2</sup>)<sup>13</sup> | 27.34 (6.31) | 27.41 (6.22) | 27.14 (6.57) | 0.983<sup>5</sup> |
| BMI categories (cut-off 30)<sup>13</sup> |  |  |  | 0.295<sup>8</sup> |
| No obesity | 210 (70.7%) | 155 (72.4%)<br>[65.8-78.2] | 55 (66.3%)<br>[55.0-76.0] |  |
| Obesity | 87 (29.3%) | 59 (27.6%)<br>[21.8-34.2] | 28 (33.7%)<br>[24.0-45.0] |  |

<sup>1</sup>Depressed if HAMD total score > 18; <sup>2</sup>For categorical variables: n (%) [95% CI, Wilson score method]. Continuous variables: mean <sup>4</sup>HAM-D: Hamilton depression rating scale; <sup>5</sup>Wilcoxon rank sum test; <sup>6</sup>GAD-7: generalised anxiety disorder assessment; <sup>7</sup>LTE: list of threatening experiences; <sup>8</sup>Pearson's Chi-squared test; <sup>9</sup>CRP: C-reactive protein (mg/L); <sup>10</sup>HDL: high-density lipoprotein (mmol/L); <sup>11</sup>LDL: low-density lipoprotein (mmol/L); <sup>12</sup>HbA1c: glycated haemoglobin (mmol/mol); <sup>13</sup>BMI: body mass index (kg/m<sup>2</sup>); CI = Confidence Interval

### Does moderate/severe depression predict poorer survival in individuals in haemodialysis?

A total of 285 participants were included in the survival analysis, of whom 111 (38.9%, 95% CI 33.5 to 44.7) died during a median follow-up of 43.3 months (IQR 22.9 to 47.1). Twelve participants (4.0%) were excluded because their vital status could not be determined: missing modality information (n = 2), loss to follow-up (n = 1), or transfer out of the haemodialysis clinic (n = 9). Kaplan–Meier analysis showed no statistically significant difference in unadjusted survival between participants with moderate/severe depressive symptoms and those with no/mild symptoms (log-rank p = 0.097, Supplementary Figure S1).

Cox proportional-hazards regression was subsequently used to estimate the association between depressive-symptom severity and mortality while accounting for follow-up time and censoring, and covariates. In models pooled across 20 multiply imputed datasets, participants with moderate/severe depressive symptoms had an estimated 49% higher mortality hazard than those with no/mild symptoms after adjustment for sex, age, ethnicity, CKD type and smoking status. However, the confidence interval included the null value, and the evidence did not meet the conventional threshold for statistical significance (HR = 1.49, 95% CI 0.99 to 2.24, p = 0.055, Figure 1B).

**Figure 1:**
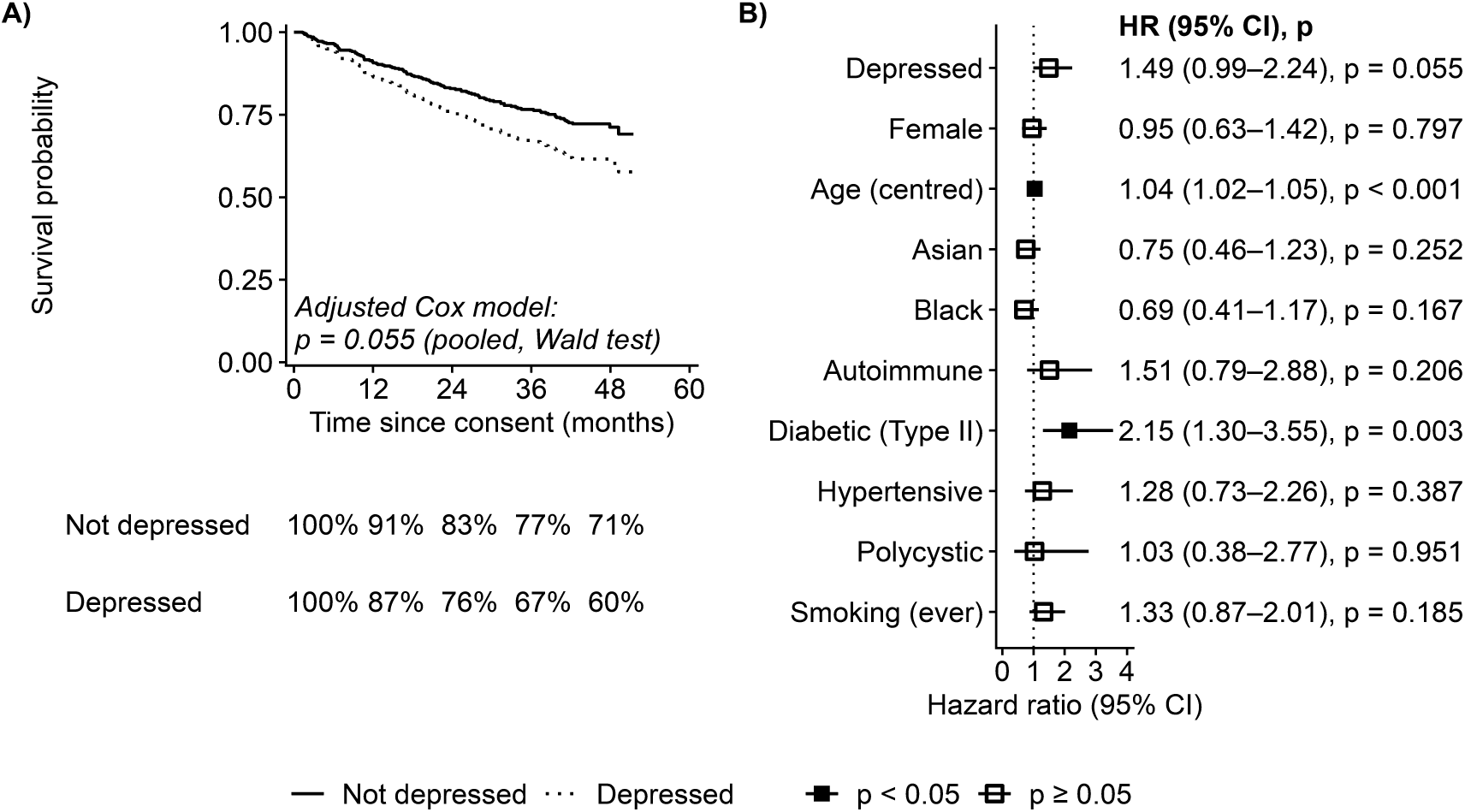
Survival analysis of CKD individuals by depressive symptoms status after 4-years follow-up, adjusted for age, gender, ethnicity, smoking and CKD subtype.

The complete-case sensitivity analysis produced a similar estimate (HR = 1.49, 95% CI 1.00 to 2.23, p = 0.052; Supplementary Table S2), supporting the direction and magnitude of the primary result but likewise providing borderline statistical evidence. The global Schoenfeld-residual test showed no evidence that the proportional-hazards assumption had been violated (p = 0.353; Supplementary Table S5).

Age was associated with mortality, with each additional year corresponding to a 4% higher mortality hazard (HR = 1.04, 95% CI 1.02 to 1.05; p < 0.001). Neither sex nor ethnicity was significantly associated with mortality in the adjusted model. Compared with CKD of uncertain aetiology, CKD attributed to type 2 diabetic nephropathy was associated with more than twice the mortality hazard (HR = 2.15, 95% CI 1.30 to 3.55, p = 0.003).

Additional adjustment for BMI produced an estimate similar in direction and magnitude to the primary adjusted estimate, although the confidence interval included the null value (HR = 1.48, 95% CI 0.98 to 2.23, p = 0.060). The association between BMI and mortality was also examined in a supplementary analysis using a BMI cut-off of 30 kg/m² (Supplementary Table S3).

### Is stress-response dysregulation associated with depressive symptoms in people receiving haemodialysis?

We first investigated whether candidate inflammatory, metabolic and cardiovascular markers related to stress-response dysregulation were associated with depressive-symptom severity, measured continuously using the HAMD-17 total score (Figure 2). We then explored biomarker-by-sex interaction terms to determine whether these associations differed between women and men.

**Figure 2:**
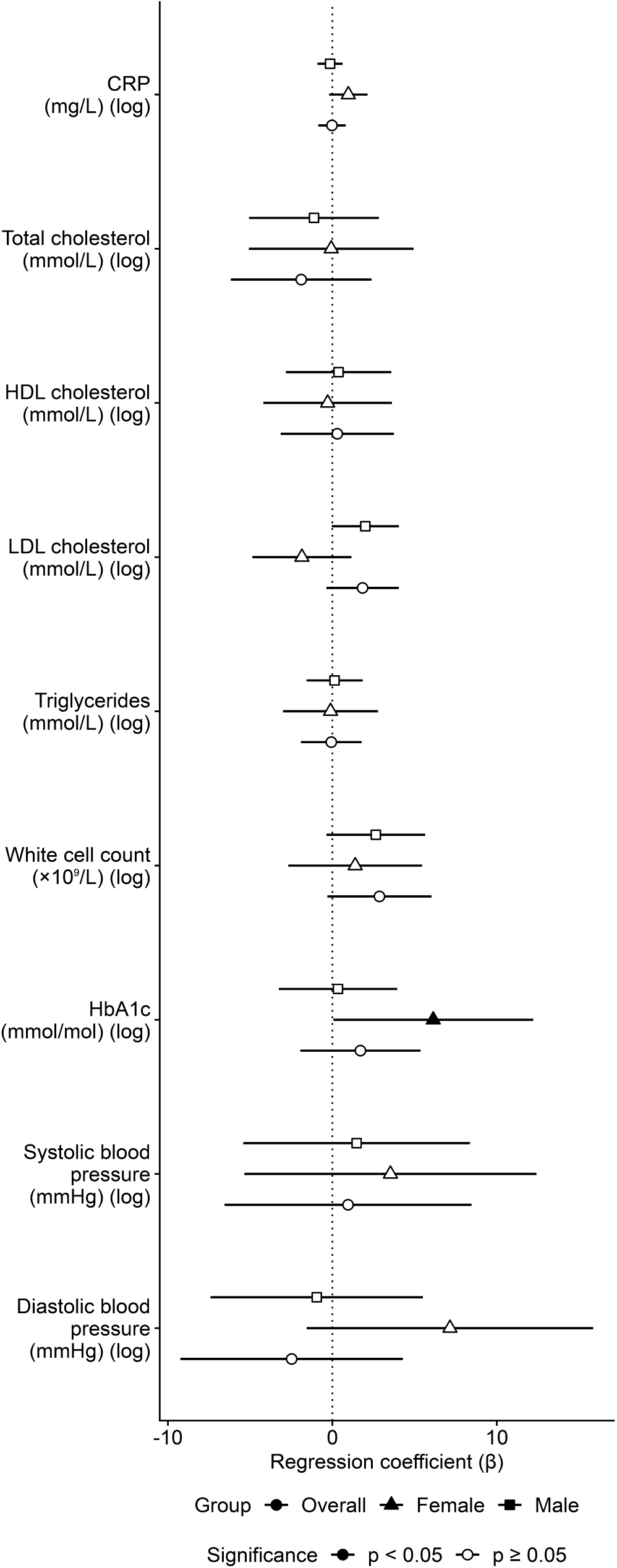
A) Stress response biomarkers association by HAMD depression total score and by gender.

### Associations with depressive symptoms

In the adjusted overall-cohort analyses, higher white-cell count was associated with greater depressive-symptom severity (β = 2.41, SE = 1.19, p = 0.044). This association was slightly attenuated after adjusting for BMI and no longer met the conventional threshold for statistical significance (β = 2.24, SE = 1.22, p = 0.068).

There was also weak evidence of a positive association between HbA1c and HAMD-17 scores in the adjusted model (β = 2.63, SE = 1.52, p = 0.086), but this was further attenuated when adjusted for BMI (β = 2.38, SE = 1.61, p = 0.140). CRP, total cholesterol, HDL cholesterol, LDL cholesterol, triglycerides, systolic blood pressure and diastolic blood pressure were not associated with depressive-symptom severity in either the adjusted or fully adjusted overall-cohort models.

### Sex differences in biomarker–depression associations

Biomarker-by-sex interaction analyses showed that diastolic blood pressure was the only marker for which there was clear evidence that the association with depressive-symptom severity differed by sex. The diastolic-blood-pressure-by-sex interaction was significant in the adjusted model (β = 11.16, SE = 4.82, p = 0.021) and remained significant after full adjustment (β = 11.14, SE = 4.82, p = 0.022; Table 1). The positive interaction coefficient indicates that the association between log-transformed diastolic blood pressure and HAMD- 17 scores was more positive in women than in men.

The LDL-cholesterol-by-sex interaction suggested a potentially weaker or inverse association among women than among men, although the evidence did not meet the conventional threshold for statistical significance after full adjustment (β = −3.27, SE = 1.73, p = 0.060).

There was no evidence of interactions for associations of CRP, HDL cholesterol, systolic blood pressure, total cholesterol, triglycerides or white-cell count with HAMD-17 scores (all fully adjusted interaction p ≥ 0.223).

There was a significant association between HBA1c and depressive symptom severity among women (β = −6.45, SE = 2.76, p = 0.022), which remained significant after BMI adjustment (β = 6.13, SE = 3.06, p = 0.048).

Overall, the strongest evidence of sex modification was observed for diastolic blood pressure.

### Do inflammatory or metabolic markers account for the association between depression status and mortality?

Exploratory models were used to examine whether stress-response biomarkers accounted for the association between depression status and mortality. Specifically, based on significance in Table S6, we examined HbA1c, WCC, and LDL cholesterol as candidate biological markers that could partly explain the depression–mortality near association observed in the primary Cox survival analysis. These analyses were considered exploratory rather than formal causal mediation models, because the observational design and timing of biomarker assessment limit inference about temporal ordering.

The fully adjusted depression-mortality total effect was significant (OR = 1.85, p = 0.048). In these exploratory models, HbA1c was not independently associated with mortality, but was close to significance and had a high odds ratio (OR = 2.54, p = 0.063) (Table S7). HbA1c had the highest depression status effect decrease when adjusted for, with a percentage change of - 6%, but did not reach significance (OR = 1.78, p = 0.066) (Table S7). Surprisingly, HbA1c was independently significant in the model with WCC and LDL cholesterol (OR = 2.80, p = 0.048) (Table S7).

WCC was not significantly associated with mortality when included with depression, either alone or in the combined model with HbA1c and LDL cholesterol (Table S7). Depression status was significantly associated with WCC for mortality (OR = 1.86, p = 0.046) and in the combined model (Table 2).

**Table 2:** Attenuation of depression status effect when adjusting for mediators.

| Model | Coefficient | SE | OR | 95% CI | p-value | % change |
| --- | --- | --- | --- | --- | --- | --- |
| Total effect (no mediators) | 0.614 | 0.310 | 1.85 | (1.01–3.39) | **0.048** | 0% |
| Adjusted for HbA1c | 0.577 | 0.312 | 1.78 | (0.97–3.28) | 0.066 | -6% |
| Adjusted for WCC | 0.623 | 0.311 | 1.86 | (1.01–3.43) | **0.046** | 1.5% |
| Adjusted for LDL cholesterol | 0.623 | 0.310 | 1.87 | (1.02–3.43) | **0.046** | 1.6% |
| Adjusted for WCC and LDL cholesterol | 0.634 | 0.311 | 1.88 | (1.02–3.47) | **0.043** | 3.3% |
| Adjusted for HbA1c, WCC and LDL cholesterol | 0.603 | 0.314 | 1.83 | (0.99–3.38) | 0.056 | -1.8% |

**Table 3:** Fully adjusted multivariate linear regression of CTRA related scores and biomarkers.

| Exposure | $\beta$ | SE | p-value |
| --- | --- | --- | --- |
| E-selectin (ng/ml) | -0.046 | 0.048 | 0.351 |
| TNFα (pg/ml)<sup>1</sup> | 0.023 | 0.013 | 0.092 |
| IL1Rα (pg/ml)<sup>2</sup> | 0.002 | 0.004 | 0.709 |
| GFAP (pg/ml)<sup>3</sup> | -0.003 | 0.012 | 0.830 |
| Inflammatory genes (Z-Score) | 0.765 | 0.938 | 0.423 |
| Type 1 Interferon (Z-Score) | 0.596 | 0.434 | 0.183 |
| CTRA (Z-Score)<sup>4</sup> | -0.459 | 0.459 | 0.328 |

LDL cholesterol was not significantly associated with mortality when included with depression status, either alone or in the combined model with HbA1c and WCC (Table S7). Depression status was significantly associated with LDL cholesterol as a mediator for mortality (OR = 1.87, p = 0.046) (Table S7) and in the combined model (Table 2).

Depression was significant in the model with both LDL and WCC as potential mediators (OR 1.88, p = 0.043) with a percent change of 3.3%. The model containing all 3 mediators did not reach significance on depression status effect (Table 2).

Depression was significantly associated with WCC and LDL cholesterol, but not with HbA1c, in the candidate mediator-path models. The findings provided preliminary evidence that WCC and LDL cholesterol may partially mediate the association between depression and mortality.

### Laboratory sub-study of stress-related transcriptomics

In a smaller laboratory sub-study, we examined whether depressive symptoms were associated with inflammatory cytokines, markers of the blood brain barrier disruption, the CTRA among individuals receiving haemodialysis. We explored whether any association was driven by specific components of the CTRA profile, namely upregulation of pro- inflammatory gene expression or downregulation of type-I interferon (IFN) related antiviral gene expression or by inflammatory cytokines levels overall and by gender. We found no evidence that deeper inflammatory profiling in this small cohort could capture further dysregulation of the stress response system.

## Discussion

This study investigated whether moderate/severe depressive symptoms in patients receiving haemodialysis were associated with mortality, stress response biomarkers, and stress-related transcriptional profiles. Moderate/severe depressive symptoms were associated with a poorer survival over follow-up. White blood cell count showed evidence of association with depression. Sex differences were observed particularly for diastolic blood pressure. Exploratory models suggested that WCC and LDL may be independently associated with depression and mortality. We found no evidence that other pro-inflammatory cytokines, overall CTRA composite or IFN-related antiviral gene expression were associated with depression in haemodialysis. Together, these findings support the view that depression in CKD may identify a clinically relevant psychobiological risk state.

We have shown that depression is an independent predictor of increased all-cause mortality in chronic kidney disease. Moderate/severe depression in haemodialysis populations is frequently conceptualised as a psychological response to severe chronic illness. While this is partly correct, our findings suggest that moderate/severe depressive symptoms may also index a broader psychobiological risk state in this population. The overlap between depression, inflammatory activity, and mortality is consistent with models in which affective symptoms, immune activation, and physical disease progression are dynamically linked rather than operating as independent clinical domains. We ourselves have previously shown using large multi-modal data that CRP mediates the association between depressive symptoms and mortality on another large dataset ^36^. The association between depressive symptoms and inflammatory markers is consistent with a substantial literature implicating immune activation in depression without physical illness ^37^. In CKD, this relationship may be particularly important because patients are already exposed to persistent inflammatory stimuli, including uraemia, oxidative stress, haemodialysis-related immune activation, vascular access complications, and high cardiometabolic burden ^38^. Our study agreed with others showing the extensive evidence between depression and type 2 diabetes, from mendelian randomisation ^39^, epidemiological ^40^ and genetic studies ^41^. Rather than indicating that inflammation is unique to depression, the present findings suggest that depressive symptoms may identify a subgroup of haemodialysis patients in whom inflammatory and glycaemic disturbances are clinically important.

The study also has implications for understanding health inequalities in CKD. The haemodialysis population served by Barts Health includes communities exposed to high levels of socioeconomic adversity and structural disadvantage. Social economic, health events and financial struggles are particular social stressors and further disadvantage this population, and cumulative life events may contribute to allostatic load and immune dysregulation. In this context, immune-metabolic disturbances in depression may represent one pathway through which adverse social environments become biologically embedded and contribute to poorer physical health outcomes. Future studies should use longitudinal designs with repeated measurement of depressive symptoms, biological and social adversity. This would allow testing of whether social stressors precede immune activation, whether inflammatory profiles predict subsequent depression or mortality, and whether depressive symptoms improve when inflammatory or social risk factors are addressed. Multi-marker immune and metabolic profiling may be particularly useful for distinguishing immune-metabolic depression from depressive symptoms driven primarily by uraemia, fatigue, or pain. Our study also agrees with others which pointed towards broader gene expression profiling of immune-related genes as a more sensitive pro-inflammatory marker for depression showing immune alterations beyond commonly measured CRP or WCC ^42^.

Moderate/severe depressive symptoms were more common among women in this cohort, consistent with the broader epidemiology of depression. This difference may reflect sex-related variation in immune regulation, susceptibility to autoimmune disease, endocrine function, and exposure to social stressors. The cohort’s mean age was approximately 60 years, by which time most women would be postmenopausal. Reproductive history and cumulative lifetime oestrogen exposure may plausibly influence vascular and immune-related ageing; however, these factors were not measured and should be investigated directly in future studies.

This study has several strengths, including recruitment across multiple haemodialysis sites, inclusion of participants from diverse ethnic backgrounds in one of the most socioeconomically deprived regions of the UK, and availability of four-year follow-up data for examining survival in relation to depressive symptoms. Several limitations should nevertheless be acknowledged. Biomarkers were measured at a single time point, and most biological comparisons were cross-sectional, precluding conclusions about causality, temporal direction or mediation. COVID-19-related suspension of research substantially delayed recruitment and data collection for the exploratory laboratory sub-study, resulting in laboratory measurements being available for only a small subset of the parent cohort and reducing statistical power and generalisability. Because follow-up also overlapped with a period of elevated mortality among people receiving haemodialysis, the laboratory subgroup may have differed systematically from participants who were not recruited or who died before laboratory sampling, introducing potential selection and survivor bias. Accordingly, the candidate immune–metabolic and transcriptional findings should be considered exploratory and require validation in larger, prospectively sampled cohorts.

In conclusion, depressive symptoms in haemodialysis patients appear to be clinically and biologically meaningful. The findings support a model in which depression, inflammation, social adversity, and survival might be interconnected in CKD. Although causality cannot be inferred, the results suggest that depression in haemodialysis populations should be approached as a psychobiological risk state, warranting integrated psychiatric, renal, immune-metabolic and social assessment. This framework may be particularly important for addressing health inequalities in ethnically diverse and socioeconomically disadvantaged CKD populations.

## Data Availability

All data produced in the present study are available upon reasonable request to the authors

## Acknowledgements

Barts Health NHS Trust and Barts Charity for their generous support for this project [G001414]. We acknowledge BCI infrastructural support from the CRUK City of London Major Centre (CTRQQR-2021\100004).

## Author Contributions

LAC: Original grant CO-I, Conceptualisation, Methodology, Writing – Original Draft Reviewing & Editing

CD, OD: Data Analysis and Writing

CC: Patient Recruitment and Questionnaires and Analysis

KB, MY: Conceptualisation, Original grant CO-I, Review and Editing

GS: Patient Recruitment, Data Collection and Analysis

SR: Patient Recruitment

GMH: Qualitative data oversight

BG: Lived experience advisor

FL: Policy Director of Kidney Care UK

SK: Clinical oversight

DR: NHS Data Linkage

KM: Clinical oversight

SC: Data analysis

All authors read and approved the final version of the paper.

