## Supplementary Material for "Depression, immune–metabolic alterations and mortality in haemodialysis: a prospective cohort study"

### KI Reports Manuscript Submission Style Checklist

#### Manuscript Formatting Checklist

◙ Make abstract structured, with introduction, methods, results and conclusion subheads.

◙ Below the title, provide corresponding author's name, e-mail address complete mailing address.

◙ Methods section must appear before results section.

◙ Include the subhead "Disclosures" in the manuscript after the discussion but before the references. Below it, list any author financial disclosures. If none, write "nothing to disclose."

◙ Provide figures as separate files in ppt, eps, jpg or tiff format. (NO PDF/png format). Place figure legends in the manuscript after the references but exclude figures from the manuscript file.

◙ Label figure files according to their number (i.e., Figure 1, etc.). Provide figures in color if possible.

◙ Place tables in the main manuscript file after the references. Create tables using MS Word.

◙ Clearly cite all tables and figures in the manuscript text with the table/figure number(s).

#### Supplemental Material

◙ COMBINE ALL SUPPLEMENTARY MATERIAL IN ONE PDF FILE, including checklist if applicable. Include all supplementary titles/legends in the supplementary file.

◙ Include subhead "Supplementary Material" in the manuscript before the references. Below, list a short title/legend for each supplementary item, the file format in parentheses [e.g., "(PDF)"], and write "Supplementary information is available at KI Report's website."

◙ Cite each individual supplementary material item in the manuscript text using the "S" prefix (e.g.,Supplementary Table S1, Supplementary Methods, Supplementary References, etc.).

THANK YOU FOR PROVIDING A CORRECTLY FORMATTED MANUSCRIPT

### Supplementary Methods

#### Post-modelling diagnostics

Influential observations were identified using Cook's distance, leverage, and DFBETA statistics; results were compared before and after removal of the most influential observations to assess robustness of the findings.

#### Model comparison

Complete-case and imputed model estimates were compared graphically and numerically to evaluate the impact of imputation on effect estimates and standard errors.

#### Colours

Colours were selected from the Okabe-Ito and Paul Tol palettes for accessibility to readers with colour vision deficiency. Colour assignment to sex categories is arbitrary and carries no implied hierarchy.

#### Conserved Transcriptional Response to Adversity (CTRA)

Tempus tubes were collected from 42 individuals receiving maintenance dialysis of which 11 had depression. RNA was isolated from tempus tubes according to manufacture protocol. RNA sequencing was performed using QuantSeq 3' FWD by lexogen. Raw reads were assessed for quality using FastQC and adapter sequences were moved using CutAdapt (done by Lexogen). Trimmed reads were aligned to the GrCh38 genome using STAR (version) v. Lowly expressed genes (sum of <10 counts across all samples) were filtered from the raw gene count matrix and normalized using variance-stabilizing transformation (VST) implemented in DESeq2 (v1.46.0). Differential gene expression analysis was conducted with DESeq2. Principal component analysis (PCA; PCAtools v2.18.0) and sample-to-sample correlation clustering on VST counts were performed to identify outlier samples.

Our CTRA analysis followed previous research (Cole et al. 2020) in assessing the CTRA profile using an a priori-defined contrast among gene expression values for 19 pro-inflammatory genes (IL1A, IL1B, IL6, IL8, TNF,PTGS1, PTGS2, FOS, FOSB, FOSL1, FOSL2, JUN, JUNB, JUND, NFKB1, NFKB2, REL, RELA, RELB), 28 genes involved in Type I interferon response (GBP1, IFI16, IFI27, IFI27L1-2, IFI30, IFI35, IFI44, IFI44L, IFI6, IFIH1, IFIT1-3, IFIT5,IFIT1L, IFITM1-3, IFITM4P, IFITM5, IFNB1, IRF2, IRF7-8, MX1-2, OAS1-3, OASL), and 3 genes involved in antibody synthesis (IGJ, IGLL1, IGLL3), with signs reversed for the antiviral and antibody-related gene sets to reflect their inverse relationship to the CTRA profile using previously described methods (Cole 2019; Cole et al. 2020). We tested the overall CTRA composite score, the pro-inflammatory and antiviral gene sets separately to determine whether subcomponents might best contribute to depression.

In short, the CTRA score was computed as the difference in pro-inflammatory score and anti-viral Z-score.

**Table S0:** R packages used in the analysis

Factors associated with depression in dialysis patients — generated 2026-06-30

| Package | Version |
|---|---|
| Analytical packages | |
| CMAverse | 0.1.0 |
| broom | 1.0.13 |
| broom.helpers | 1.22.0 |
| car | 3.1.5 |
| cowplot | 1.2.0 |
| data.table | 1.18.4 |
| dplyr | 1.2.1 |
| easystats | 0.7.6 |
| flextable | 0.10.0 |
| forcats | 1.0.1 |
| formattable | 0.2.1 |
| ggmice | 0.1.1 |
| ggplot2 | 4.0.3 |
| ggpubr | 1.0.0 |
| ggrepel | 0.9.8 |
| ggstatsplot | 1.0.0 |
| ggtext | 0.1.2 |
| gt | 1.3.0 |
| gtExtras | 0.6.2 |
| gtsummary | 2.5.1 |
| interactions | 1.2.0 |
| janitor | 2.2.1 |
| labelled | 2.16.0 |
| magick | 2.9.1 |
| mgcv | 1.9.4 |
| mice | 3.19.0 |
| officer | 0.7.6 |
| openxlsx2 | 1.28 |
| pROC | 1.19.0.1 |

| Package | Version |
|---|---|
| parameters | 0.29.2 |
| patchwork | 1.3.2 |
| performance | 0.17.1 |
| prismatic | 1.1.2 |
| purrr | 1.2.2 |
| readxl | 1.5.0 |
| report | 0.6.4 |
| scales | 1.4.0 |
| see | 0.14.1 |
| sjlabelled | 1.2.0 |
| sjmisc | 2.8.11 |
| skimr | 2.2.2 |
| stringr | 1.6.0 |
| survival | 3.8.9 |
| survminer | 0.5.2 |
| writexl | 1.5.4 |
| Infrastructure / reproducibility | |
| fs | 2.1.0 |
| glue | 1.8.1 |
| grateful | 0.3.0 |
| here | 1.0.2 |
| knitr | 1.51 |
| quarto | 1.5.1 |
| ragg | 1.5.2 |
| renv | 1.2.3 |
| rmarkdown | 2.31 |
| sessioninfo | 1.2.4 |
| svglite | 2.2.2 |
| systemfonts | 1.3.2 |

Analyses were run in R 4.6.1. Package citations were generated with the grateful package. Software citations are distinct from the methodological references (e.g. the mediation decomposition and multiple-imputation method papers), which are cited in the main text.
Generated by report_04_cite_packages.R on 2026-07-23 17:12.

**Table S1a**: Social, demographic and clinical characteristics of individuals with CKD by depression status[1] complete case analysis.

| **Characteristic**[2] | **Overall**<br>N = 297 | **Not depressed**<br>N = 214 | **Depressed**<br>N = 83 | **p-value** |
|---|---|---|---|---|
| HAMD total score[3] | 13.58 (7.63) | 9.71 (4.57) | 23.54 (4.06) | **<0.001**[4] |
| GAD total score[5] | 3.79 (4.82) | 2.21 (3.54) | 7.87 (5.30) | **<0.001**[4] |
| LTE total score[6] | 1.69 (1.20) | 1.57 (1.13) | 1.98 (1.32) | **0.006**[4] |
| Survival status | | | | 0.104[7] |
| Alive | 174 (100.0%) | 130 (74.7%) [67.5-80.8] | 44 (25.3%) [19.2-32.5] | |
| Deceased | 111 (100.0%) | 73 (65.8%) [56.1-74.3] | 38 (34.2%) [25.7-43.9] | |
| NA | 12 | 11 | 1 | |
| Gender | | | | **0.012**[7] |
| Female | 123 (100.0%) | 79 (64.2%) [55.0-72.5] | 44 (35.8%) [27.5-45.0] | |
| Male | 174 (100.0%) | 135 (77.6%) [70.5-83.4] | 39 (22.4%) [16.6-29.5] | |
| Age | 60.39 (15.18) | 61.21 (14.90) | 58.30 (15.78) | 0.186[4] |
| High socio-economic status | 56 (100.0%) | 44 (78.6%) [65.2-88.0] | 12 (21.4%) [12.0-34.8] | 0.231[7] |
| NA | 3 | 2 | 1 | |
| Married status | 160 (100.0%) | 113 (70.6%) [62.8-77.4] | 47 (29.4%) [22.6-37.2] | 0.553[7] |
| Ethnicity | | | | 0.564[7] |
| Asian | 108 (100.0%) | 74 (68.5%) [58.8-76.9] | 34 (31.5%) [23.1-41.2] | |
| Black | 100 (100.0%) | 75 (75.0%) [65.2-82.9] | 25 (25.0%) [17.1-34.8] | |
| White | 89 (100.0%) | 65 (73.0%) [62.4-81.6] | 24 (27.0%) [18.4-37.6] | |
| Chronic kidney disease type | | | | 0.904[7] |
| Autoimmune | 45 (100.0%) | 33 (73.3%) [57.8-84.9] | 12 (26.7%) [15.1-42.2] | |
| Diabetic (Type II) | 64 (100.0%) | 43 (67.2%) [54.2-78.1] | 21 (32.8%) [21.9-45.8] | |
| Hypertensive | 67 (100.0%) | 50 (74.6%) [62.3-84.1] | 17 (25.4%) [15.9-37.7] | |
| Polycystic | 18 (100.0%) | 13 (72.2%) [46.4-89.3] | 5 (27.8%) [10.7-53.6] | |
| Uncertain | 103 (100.0%) | 75 (72.8%) [63.0-80.9] | 28 (27.2%) [19.1-37.0] | |
| Creatinine (µmol/L) | 774.35 (299.88) | 780.63 (290.96) | 758.31 (322.84) | 0.428[4] |
| NA | 2 | 2 | 0 | |

| Characteristic[2] | Overall<br>N = 297 | Not depressed<br>N = 214 | Depressed<br>N = 83 | p-value |
|---|---|---|---|---|
| eGFR (ml/min/1.73m²)[8] | 7.72 (8.66) | 7.74 (9.32) | 7.66 (6.76) | 0.712[4] |
| NA | 3 | 3 | 0 | |
| Cardiovascular disease | 114 (100.0%) | 79 (69.3%) [59.9-77.4] | 35 (30.7%) [22.6-40.1] | 0.404[7] |
| Hypertension | 174 (100.0%) | 126 (72.4%) [65.0-78.8] | 48 (27.6%) [21.2-35.0] | 0.869[7] |
| Systolic blood pressure | 149.03 (23.81) | 147.47 (23.31) | 153.13 (24.77) | 0.074[4] |
| NA | 1 | 0 | 1 | |
| Diastolic blood pressure | 76.77 (14.29) | 75.87 (13.30) | 79.10 (16.48) | 0.305[4] |
| NA | 1 | 0 | 1 | |
| Mean arterial blood pressure (mmHg) | 100.86 (16.00) | 99.74 (15.27) | 103.78 (17.55) | 0.122[4] |
| NA | 1 | 0 | 1 | |
| Diabetes (based on CKD)[9] | | | | >0.999[10] |
| Diabetes (Type I) | 11 (100.0%) | 8 (72.7%) [39.3-92.7] | 3 (27.3%) [7.33-60.7] | |
| Diabetes (Type II) | 64 (100.0%) | 43 (67.2%) [54.2-78.1] | 21 (32.8%) [21.9-45.8] | |
| NA | 222 | 163 | 59 | |
| Smoking status | | | | 0.220[7] |
| No | 197 (100.0%) | 148 (75.1%) [68.4-80.9] | 49 (24.9%) [19.1-31.6] | |
| No, but used to | 68 (100.0%) | 46 (67.6%) [55.1-78.2] | 22 (32.4%) [21.8-44.9] | |
| Yes | 32 (100.0%) | 20 (62.5%) [43.7-78.3] | 12 (37.5%) [21.7-56.3] | |
| Smoking (ever) | 100 (100.0%) | 66 (66.0%) [55.8-75.0] | 34 (34.0%) [25.0-44.2] | 0.098[7] |
| Smoking (currently) | 32 (100.0%) | 20 (62.5%) [43.7-78.3] | 12 (37.5%) [21.7-56.3] | 0.202[7] |
| BMI (kg/m²)[11] | 27.38 (6.78) | 27.48 (6.67) | 27.12 (7.08) | 0.842[4] |
| NA | 49 | 35 | 14 | |
| BMI categories (cut-off 30)[11] | | | | 0.257[7] |
| No obesity | 168 (100.0%) | 125 (74.4%) [67.0-80.7] | 43 (25.6%) [19.3-33.0] | |
| Obesity | 80 (100.0%) | 54 (67.5%) [56.0-77.3] | 26 (32.5%) [22.7-44.0] | |
| NA | 49 | 35 | 14 | |

[1]Depressed if HAMD total score > 18

[2]For categorical variables: n (%) [95% CI, Wilson score method]. Continuous variables: mean (SD).

[3]HAM-D: Hamilton depression rating scale

[4]Wilcoxon rank sum test

[5]GAD-7: generalised anxiety disorder assessment

[6]LTE: list of threatening experiences

[7]Pearson's Chi-squared test

[8]eGFR: estimated glomerular filtration rate (ml/min/1.73m²)

[9]CKD: Chronic kidney disease type

[10]Fisher's exact test

[11]BMI: body mass index (kg/m²)

Abbreviation: CI = Confidence Interval

**Table S2**: Biomarkers of stress response by depression status [1] complete case analysis.

| Characteristic | **Not depressed** N = 214[2] | **Depressed** N = 83[2] | **p-value** |
|---|---|---|---|
| CRP (mg/L)[3] | 10.98 (16.96) | 14.17 (20.56) | 0.113[4] |
| NA | 2 | 0 | |
| Total cholesterol (mmol/L) | 3.85 (1.14) | 4.00 (1.18) | 0.307[4] |
| NA | 3 | 0 | |
| HDL cholesterol (mmol/L)[5] | 1.24 (0.49) | 1.21 (0.41) | 0.938[4] |
| NA | 29 | 13 | |
| LDL cholesterol (mmol/L)[6] | 2.04 (0.98) | 2.04 (0.97) | 0.912[4] |
| NA | 31 | 12 | |
| Triglycerides (mmol/L) | 1.77 (1.48) | 2.11 (2.25) | 0.669[4] |
| NA | 29 | 13 | |
| White cell count (×10⁹/L) | 6.86 (2.59) | 7.68 (3.23) | 0.087[4] |
| NA | 1 | 0 | |
| HbA1c (mmol/mol)[7] | 43.14 (16.51) | 45.92 (17.22) | 0.155[4] |
| NA | 10 | 7 | |

[1]Depressed if HAMD total score > 18

[2]Mean (SD)

[3]CRP: C-reactive protein (mg/L)

[4]Wilcoxon rank sum test

[5]HDL: high-density lipoprotein (mmol/L)

[6]LDL: low-density lipoprotein (mmol/L)

[7]HbA1c: glycated haemoglobin (mmol/mol)

**Figure S1** A): Kaplan-Meier plot of survival by depression status over a 48-months follow up period; B) Cox proportional hazards regression: hazard ratios for mortality (complete-case data)

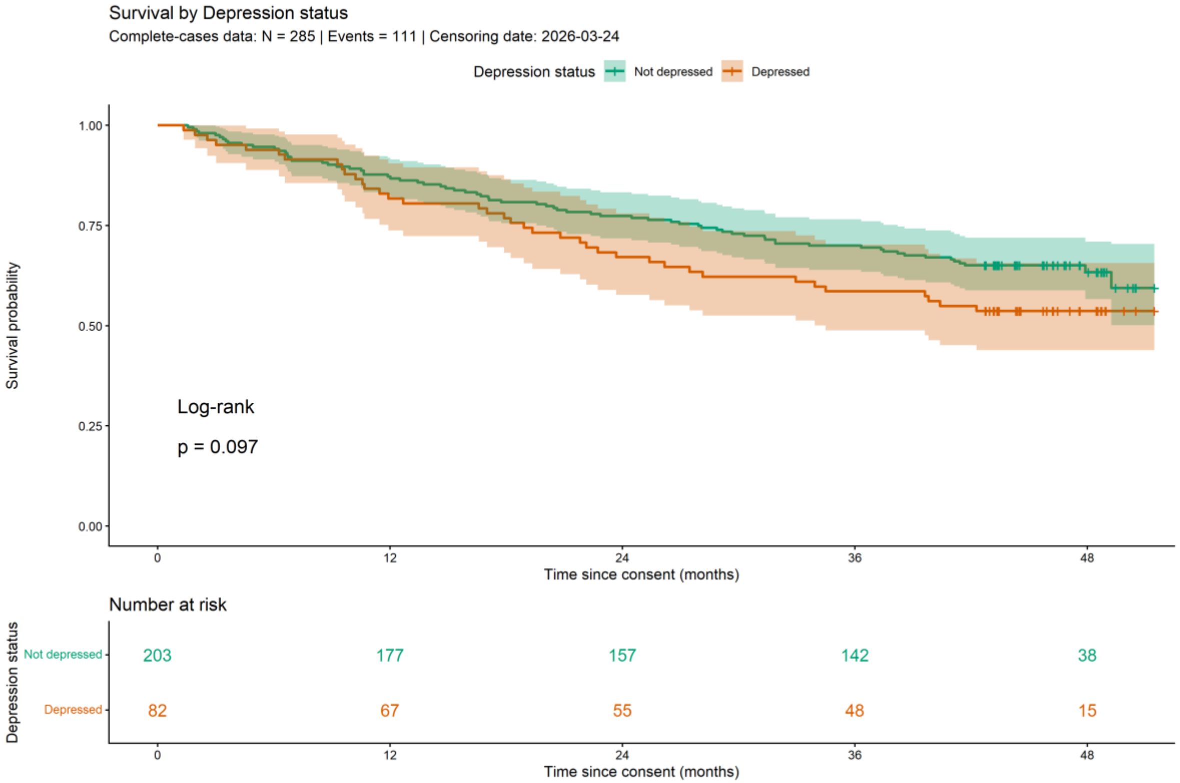

| | Adjusted | | |
|---|---|---|---|
| **Characteristic** | **HR** | **95% CI** | **p-value** |
| Depression status | | | |
| Not depressed (Ref.) | | | |
| Depressed | **1.49** | **1.00, 2.23** | **0.052** |
| Gender | | | |
| Male (Ref.) | | | |
| Female | 0.95 | 0.64, 1.42 | 0.797 |
| Age (centred) | 1.04 | 1.02, 1.05 | **<0.001** |
| Ethnicity | | | |
| White (Ref.) | | | |
| Asian | 0.75 | 0.47, 1.22 | 0.249 |
| Black | 0.69 | 0.41, 1.16 | 0.164 |
| Chronic kidney disease type | | | |
| Uncertain (Ref.) | | | |
| Autoimmune | 1.51 | 0.80, 2.86 | 0.203 |
| Diabetic (Type II) | 2.15 | 1.31, 3.53 | **0.002** |
| Hypertensive | 1.28 | 0.73, 2.25 | 0.385 |
| Polycystic | 1.03 | 0.39, 2.74 | 0.951 |
| Smoking (ever) | | | |
| No (Ref.) | | | |
| Yes | 1.33 | 0.88, 2.00 | 0.182 |

Abbreviations: CI = Confidence Interval, HR = Hazard Ratio

**Table S3**: PH regression by BMI categories (cut-off 30) subgroup (pooled imputed data)

| | **Unadjusted** | | | **Adjusted** | | |
|---|---|---|---|---|---|---|
| **Characteristic** | **HR** | **95% CI** | **p-value** | **HR** | **95% CI** | **p-value** |
| **BMI categories (cut-off 30): No obesity** | | | | | | |
| Depression status | | | | | | |
| Not depressed (Ref.) | | | | | | |
| Depressed | 1.50 | 0.90, 2.51 | 0.115 | 1.59 | 0.92, 2.72 | 0.092 |
| Gender | | | | | | |
| Male (Ref.) | | | | | | |
| Female | | | | 0.91 | 0.54, 1.54 | 0.717 |
| Age (centred) | | | | 1.05 | 1.03, 1.07 | **<0.001** |
| Ethnicity | | | | | | |
| White (Ref.) | | | | | | |
| Asian | | | | 0.78 | 0.42, 1.45 | 0.432 |
| Black | | | | 0.75 | 0.37, 1.52 | 0.419 |
| Chronic kidney disease type | | | | | | |
| Uncertain (Ref.) | | | | | | |
| Autoimmune | | | | 1.54 | 0.65, 3.62 | 0.320 |
| Diabetic (Type II) | | | | 2.15 | 1.13, 4.08 | **0.020** |
| Hypertensive | | | | 1.21 | 0.59, 2.47 | 0.604 |
| Polycystic | | | | 1.21 | 0.39, 3.77 | 0.734 |
| Smoking (ever) | | | | | | |
| No (Ref.) | | | | | | |
| Yes | | | | 1.19 | 0.69, 2.03 | 0.526 |
| **BMI categories (cut-off 30): Obesity** | | | | | | |
| Depression status | | | | | | |
| Not depressed (Ref.) | | | | | | |
| Depressed | 1.16 | 0.57, 2.39 | 0.670 | 1.30 | 0.60, 2.81 | 0.493 |
| Gender | | | | | | |
| Male (Ref.) | | | | | | |
| Female | | | | 0.97 | 0.44, 2.14 | 0.937 |
| Age (centred) | | | | 1.02 | 0.99, 1.05 | 0.155 |
| Ethnicity | | | | | | |
| White (Ref.) | | | | | | |
| Asian | | | | 0.75 | 0.29, 1.95 | 0.538 |
| Black | | | | 0.62 | 0.22, 1.72 | 0.343 |
| Chronic kidney disease type | | | | | | |
| Uncertain (Ref.) | | | | | | |
| Autoimmune | | | | 1.52 | 0.44, 5.18 | 0.490 |
| Diabetic (Type II) | | | | 2.03 | 0.74, 5.59 | 0.161 |
| Hypertensive | | | | 1.24 | 0.38, 4.02 | 0.708 |
| Polycystic | | | | 0.78 | 0.08, 7.46 | 0.822 |
| Smoking (ever) | | | | | | |
| No (Ref.) | | | | | | |
| Yes | | | | 1.58 | 0.66, 3.74 | 0.288 |

HR = hazard ratio; CI = confidence interval
Abbreviations: CI = Confidence Interval, HR = Hazard Ratio

**Table S4**: Proportional hazards assumption (Schoenfeld test)

Adjusted Cox model (BMI not included; see Table 10.3b for BMI subgroup analysis)

| Variable | chisq | df | p |
|---|---|---|---|
| hamd_txt | 0.04 | 1 | 0.838 |
| gender | 0.49 | 1 | 0.482 |
| cage | 0.53 | 1 | 0.465 |
| ethnicity_cat | 0.32 | 2 | 0.854 |
| ckd3 | 7.97 | 4 | 0.093 |
| smoking_ever | 0.73 | 1 | 0.393 |
| GLOBAL | 11.06 | 10 | 0.353 |

A significant p-value (< 0.05) suggests violation of the PH assumption

GLOBAL = overall test for the entire model

**Table S5**: Multivariate linear regression (Adjusted | all BMI categories (cut-off 30) strata) - Outcome: HAMD total score

| | Overall | | | | Female | | | | Male | | | | Interaction | | |
|---|---|---|---|---|---|---|---|---|---|---|---|---|---|---|---|
| **Exposure** | **FMI** | **Beta** | **SE** | **p-value** | **FMI** | **Beta** | **SE** | **p-value** | **FMI** | **Beta** | **SE** | **p-value** | **Beta** | **SE** | **p-value** |
| **BMI categories (cut-off 30): No obesity** | | | | | | | | | | | | | | | |
| CRP (mg/L) (log)[1] | 5.6% | 0.03 | 0.47 | 0.949 | 5.0% | 0.16 | 0.71 | 0.827 | 6.1% | -0.08 | 0.44 | 0.853 | 0.09 | 0.77 | 0.903 |
| Total cholesterol (mmol/L) (log) | 5.7% | -2.03 | 2.54 | 0.424 | 12.6% | -3.26 | 3.28 | 0.326 | 7.2% | -1.31 | 2.41 | 0.588 | -0.16 | 3.82 | 0.967 |
| HDL cholesterol (mmol/L) (log)[2] | 24.0% | 0.32 | 2.12 | 0.879 | 10.4% | 0.07 | 2.38 | 0.976 | 25.4% | 0.41 | 2.00 | 0.839 | 0.14 | 3.01 | 0.963 |
| LDL cholesterol (mmol/L) (log)[3] | 10.8% | 1.70 | 1.40 | 0.226 | 8.6% | 0.12 | 1.82 | 0.948 | 11.9% | 1.87 | 1.31 | 0.157 | -1.09 | 2.09 | 0.601 |
| Triglycerides (mmol/L) (log) | 11.2% | 0.21 | 1.07 | 0.842 | 32.5% | -0.01 | 2.10 | 0.997 | 11.4% | 0.31 | 1.00 | 0.753 | 0.21 | 1.97 | 0.917 |
| White cell count (×10⁹/L) (log) | 2.9% | 2.79 | 1.76 | 0.115 | 5.0% | -1.37 | 2.59 | 0.598 | 3.4% | 2.66 | 1.70 | 0.119 | -3.90 | 2.64 | 0.142 |
| HbA1c (mmol/mol) (log)[4] | 7.4% | 1.94 | 2.20 | 0.380 | 13.6% | 6.85 | 4.25 | 0.113 | 8.8% | 0.57 | 2.15 | 0.793 | 1.61 | 3.75 | 0.669 |
| Systolic blood pressure (mmHg) (log) | 15.8% | 3.30 | 4.69 | 0.483 | 7.0% | 8.36 | 5.89 | 0.161 | 15.0% | 3.23 | 4.39 | 0.464 | 3.39 | 6.78 | 0.618 |
| Diastolic blood pressure (mmHg) (log) | 7.3% | -1.47 | 3.94 | 0.709 | 11.1% | 7.32 | 6.20 | 0.243 | 8.8% | -0.25 | 3.80 | 0.948 | 10.89 | 6.18 | 0.080 |
| **BMI categories (cut-off 30): Obesity** | | | | | | | | | | | | | | | |
| CRP (mg/L) (log)[1] | 18.3% | -0.44 | 1.16 | 0.707 | 10.9% | 3.04 | 1.31 | **0.027** | 24.6% | -0.68 | 1.09 | 0.537 | 2.31 | 1.51 | 0.129 |
| Total cholesterol (mmol/L) (log) | 12.6% | -0.62 | 4.81 | 0.897 | 10.8% | 5.21 | 4.82 | 0.287 | 16.7% | -1.38 | 4.43 | 0.757 | 5.94 | 6.26 | 0.346 |
| HDL cholesterol (mmol/L) (log)[2] | 34.0% | 0.49 | 4.50 | 0.914 | 13.0% | 2.68 | 4.16 | 0.523 | 46.0% | -0.21 | 4.46 | 0.964 | 0.05 | 5.52 | 0.993 |
| **LDL cholesterol (mmol/L) (log)[3]** | **14.1%** | **2.60** | **1.93** | **0.181** | **17.5%** | **-5.57** | **2.63** | **0.043** | **23.8%** | **2.51** | **1.88** | **0.192** | **-7.24** | **3.02** | **0.020** |
| Triglycerides (mmol/L) (log) | 17.4% | -0.85 | 2.19 | 0.698 | 17.7% | -1.18 | 2.29 | 0.610 | 25.8% | -1.01 | 2.03 | 0.624 | 0.24 | 2.86 | 0.934 |
| White cell count (×10⁹/L) (log) | 15.9% | 2.55 | 4.51 | 0.574 | 7.1% | 5.69 | 3.64 | 0.127 | 21.4% | 4.15 | 5.16 | 0.429 | 2.53 | 5.29 | 0.634 |
| HbA1c (mmol/mol) (log)[4] | 14.3% | -0.85 | 3.99 | 0.833 | 12.5% | 3.91 | 5.34 | 0.469 | 24.9% | -2.84 | 4.16 | 0.501 | 1.91 | 5.62 | 0.735 |
| Systolic blood pressure (mmHg) (log) | 17.7% | -8.38 | 8.94 | 0.352 | 12.0% | -2.55 | 8.03 | 0.753 | 25.5% | -5.45 | 8.49 | 0.527 | 5.75 | 11.57 | 0.621 |
| Diastolic blood pressure (mmHg) (log) | 17.2% | -6.57 | 8.51 | 0.443 | 16.3% | 9.98 | 7.77 | 0.208 | 21.0% | -3.48 | 8.33 | 0.679 | 16.21 | 10.50 | 0.128 |

FMI = Fraction of Missing Information; β = Regression coefficient; SE = Standard Error; p = p-value

[1]CRP: C-reactive protein (mg/L)

[2]HDL: high-density lipoprotein (mmol/L)

[3]LDL: low-density lipoprotein (mmol/L)

[4]HbA1c: glycated haemoglobin (mmol/mol)

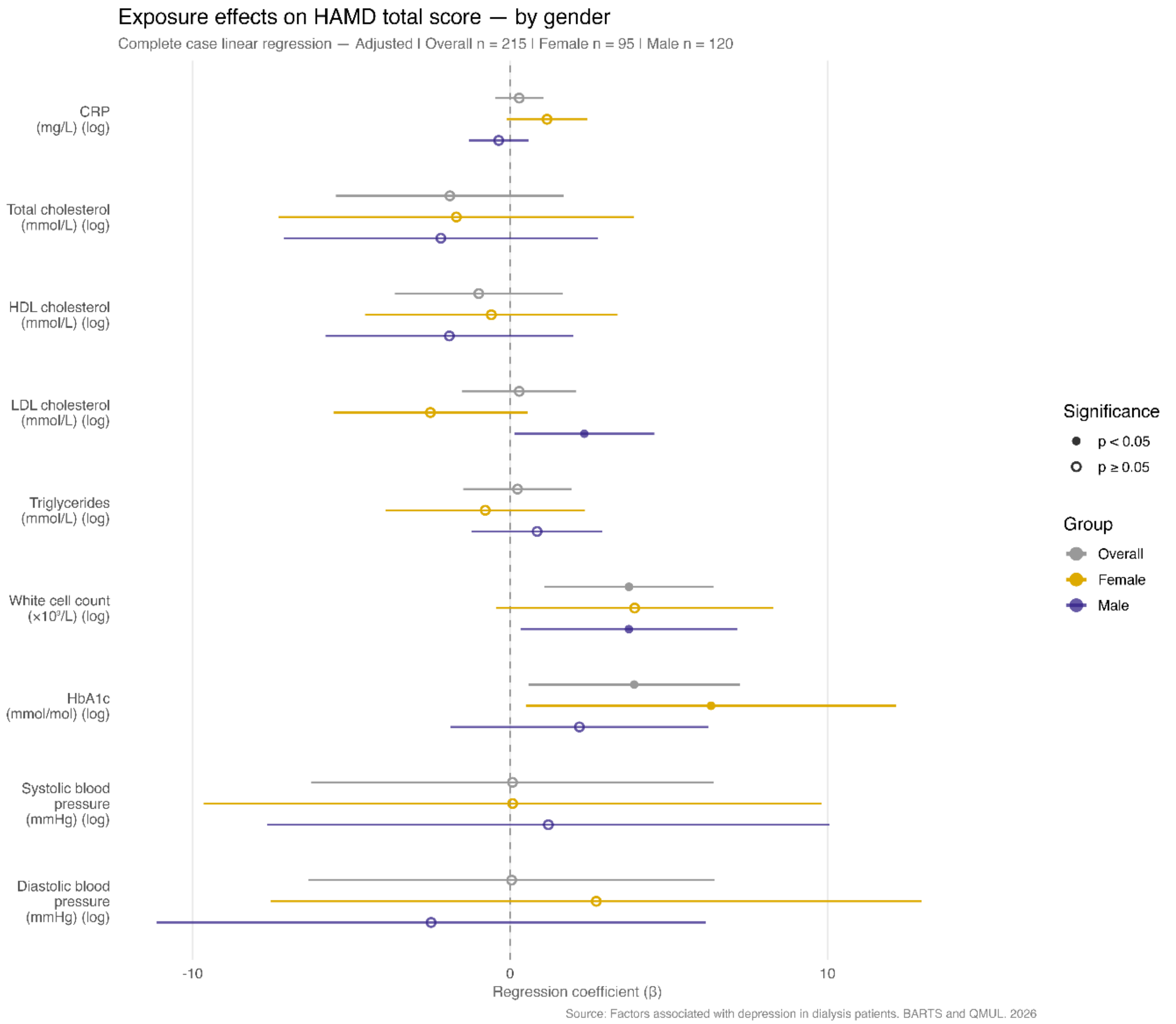

**Figure S2**: Exposure effects on depression overall and by gender (complete-case)

Outcome: HAMD total score

Fully adjusted: exposure + Gender + Age (centred) + Ethnicity + Chronic kidney disease type + Smoking (ever) + BMI (centred)

CRP: C-reactive protein (mg/L)

HDL: high-density lipoprotein (mmol/L)

LDL: low-density lipoprotein (mmol/L)

HbA1c: glycated haemoglobin (mmol/mol)

**Table S6**: Complete cases by gender - Multivariate linear regression (Adjusted) - Outcome: HAMD total score

| | Fully adjusted | | | | | | | | | | | |
|---|---|---|---|---|---|---|---|---|---|---|---|---|
| | Overall | | | Female | | | Male | | | Interaction | | |
| Exposure | β | SE | p-value | β | SE | p-value | β | SE | p-value | β | SE | p-value |
| CRP (mg/L) (log)[1] | 0.25 | 0.39 | 0.516 | 1.12 | 0.66 | 0.094 | -0.37 | 0.48 | 0.445 | 1.09 | 0.74 | 0.145 |
| Total cholesterol (mmol/L) (log) | -1.79 | 1.84 | 0.331 | -1.43 | 2.86 | 0.619 | -2.17 | 2.52 | 0.389 | 1.96 | 3.61 | 0.588 |
| HDL cholesterol (mmol/L) (log)[2] | -0.88 | 1.36 | 0.519 | -0.41 | 2.02 | 0.839 | -1.98 | 2.03 | 0.333 | 1.65 | 2.70 | 0.542 |
| LDL cholesterol (mmol/L) (log)[3] | 0.43 | 0.93 | 0.649 | **-2.38** | **1.58** | **0.137** | **2.46** | **1.14** | **0.033** | **-4.06** | **1.81** | **0.026** |
| Triglycerides (mmol/L) (log) | 0.18 | 0.87 | 0.834 | -0.82 | 1.59 | 0.607 | 0.87 | 1.07 | 0.419 | -1.27 | 1.71 | 0.458 |
| White cell count (×10⁹/L) (log) | 3.79 | 1.40 | **0.007** | 3.90 | 2.41 | 0.109 | 3.77 | 1.74 | **0.033** | -0.64 | 2.47 | 0.794 |
| HbA1c (mmol/mol) (log)[4] | 4.07 | 1.81 | **0.026** | 6.55 | 3.23 | **0.046** | 2.35 | 2.17 | 0.282 | 1.29 | 3.08 | 0.675 |
| Systolic blood pressure (mmHg) (log) | 0.12 | 3.22 | 0.970 | 0.18 | 4.91 | 0.972 | 1.22 | 4.49 | 0.787 | -1.05 | 6.31 | 0.868 |
| Diastolic blood pressure (mmHg) (log) | 0.30 | 3.28 | 0.926 | 3.21 | 5.21 | 0.539 | -2.47 | 4.42 | 0.577 | 9.16 | 5.85 | 0.119 |

β = Regression coefficient; SE = Standard Error
Fully adjusted — Female / Male: Fully adjusted: exposure + Age (centred) + Ethnicity + Chronic kidney disease type + Smoking (ever) + BMI (centred)
[1]CRP: C-reactive protein (mg/L)
[2]HDL: high-density lipoprotein (mmol/L)
[3]LDL: low-density lipoprotein (mmol/L)
[4]HbA1c: glycated haemoglobin (mmol/mol)

**Table S7**: Mediation path estimates: Depression status → HbA1c (mmol/mol), White cell count (×10⁹/L) and LDL cholesterol (mmol/L) → Mortality

| Model | Term | Estimate | SE | OR | p-value |
|---|---|---|---|---|---|
| 1. Total effect (Depression status → mortality) | hamd_txtDepressed[1] | 0.614 | 0.310 | 1.85 | **0.048** |
| 2a. Path a (Depression status → HbA1c (mmol/mol)) | hamd_txtDepressed[1] | 0.050 | 0.040 | — | 0.212 |
| 2b. Direct + HbA1c (mmol/mol) (Depression status + HbA1c (mmol/mol) → mortality) | hamd_txtDepressed[1] | 0.577 | 0.312 | 1.78 | 0.066 |
| 2b. Direct + HbA1c (mmol/mol) (Depression status + HbA1c (mmol/mol) → mortality) | log_hba1c_value | 0.931 | 0.499 | 2.54 | 0.063 |
| 3a. Path a (Depression status → White cell count (×10⁹/L)) | hamd_txtDepressed[1] | 0.061 | 0.048 | — | 0.205 |
| 3b. Direct + White cell count (×10⁹/L) (Depression status + White cell count (×10⁹/L) → mortality) | hamd_txtDepressed[1] | 0.623 | 0.311 | 1.86 | **0.046** |
| 3b. Direct + White cell count (×10⁹/L) (Depression status + White cell count (×10⁹/L) → mortality) | log_wcc_value | -0.165 | 0.395 | 0.85 | 0.677 |
| 4a. Path a (Depression status → LDL cholesterol (mmol/L)) | hamd_txtDepressed[1] | 0.027 | 0.078 | — | 0.735 |
| 4b. Direct + LDL cholesterol (mmol/L) (Depression status + LDL cholesterol (mmol/L) → mortality) | hamd_txtDepressed[1] | 0.623 | 0.310 | 1.87 | **0.046** |
| 4b. Direct + LDL cholesterol (mmol/L) (Depression status + LDL cholesterol (mmol/L) → mortality) | log_ldl_value | -0.230 | 0.306 | 0.79 | 0.455 |

| Model | Term | Estimate | SE | OR | p-value |
|---|---|---|---|---|---|
| 5. All mediators (Depression status + White cell count (×10⁹/L) and LDL cholesterol (mmol/L) → mortality) | hamd_txtDepressed[1] | 0.634 | 0.311 | 1.88 | **0.043** |
| 5. All mediators (Depression status + White cell count (×10⁹/L) and LDL cholesterol (mmol/L) → mortality) | log_wcc_value | -0.178 | 0.397 | 0.84 | 0.655 |
| 5. All mediators (Depression status + White cell count (×10⁹/L) and LDL cholesterol (mmol/L) → mortality) | log_ldl_value | -0.234 | 0.305 | 0.79 | 0.446 |
| 6. All mediators (Depression status + HbA1c (mmol/mol), White cell count (×10⁹/L) and LDL cholesterol (mmol/L) → mortality) | hamd_txtDepressed[1] | 0.603 | 0.314 | 1.83 | 0.056 |
| 6. All mediators (Depression status + HbA1c (mmol/mol), White cell count (×10⁹/L) and LDL cholesterol (mmol/L) → mortality) | log_hba1c_value | 1.029 | 0.517 | 2.8 | **0.048** |
| 6. All mediators (Depression status + HbA1c (mmol/mol), White cell count (×10⁹/L) and LDL cholesterol (mmol/L) → mortality) | log_wcc_value | -0.403 | 0.420 | 0.67 | 0.339 |
| 6. All mediators (Depression status + HbA1c (mmol/mol), White cell count (×10⁹/L) and LDL cholesterol (mmol/L) → mortality) | log_ldl_value | -0.194 | 0.304 | 0.82 | 0.525 |

All models adjusted for: Gender, Age (centred), Ethnicity, Chronic kidney disease type, Smoking (ever), BMI (centred)

OR shown for logistic models; Estimate is the regression coefficient for linear models

[1]Depression status: 'Not depressed' = HAMD total ≤ 18; 'Depressed' = HAMD total > 18

### Packages

Allaire, J. J., & Dervieux, C. (2025). *quarto: R Interface to `Quarto' Markdown Publishing System*. https://doi.org/10.32614/CRAN.package.quarto
Allaire, J. J., Xie, Y., Dervieux, C., McPherson, J., Luraschi, J., Ushey, K., Atkins, A., Wickham, H., Cheng, J., Chang, W., & Iannone, R. (2026). *rmarkdown: Dynamic Documents for R*. https://github.com/rstudio/rmarkdown
Barbone, J. M., & Garbuszus, J. M. (2026). *openxlsx2: Read, Write and Edit `xlsx' Files*. https://janmarvin.github.io/openxlsx2/
Barrett, T., Dowle, M., Srinivasan, A., Gorecki, J., Chirico, M., Hocking, T., Schwendinger, B., & Krylov, I. (2026). *data.table: Extension of `data.frame'*. https://doi.org/10.32614/CRAN.package.data.table
Buuren, S. van, & Groothuis-Oudshoorn, K. (2011). mice: Multivariate Imputation by Chained Equations in R. *Journal of Statistical Software*, *45*(3), 1–67. https://doi.org/10.18637/jss.v045.i03
Firke, S. (2024). *janitor: Simple Tools for Examining and Cleaning Dirty Data*. https://doi.org/10.32614/CRAN.package.janitor
Fox, J., & Weisberg, S. (2019). *An R Companion to Applied Regression* (Third). Sage. https://www.john-fox.ca/Companion/
Gohel, D., Moog, S., & Heckmann, M. (2026). *officer: Manipulation of Microsoft Word and PowerPoint Documents*. https://doi.org/10.32614/CRAN.package.officer
Gohel, D., & Skintzos, P. (2026). *flextable: Functions for Tabular Reporting*. https://doi.org/10.32614/CRAN.package.flextable
Hester, J., & Bryan, J. (2026). *glue: Interpreted String Literals*. https://doi.org/10.32614/CRAN.package.glue
Hester, J., Wickham, H., & Csárdi, G. (2026). *fs: Cross-Platform File System Operations Based on `libuv'*. https://doi.org/10.32614/CRAN.package.fs
Hvitfeldt, E. (2024). *prismatic: Color Manipulation Tools*. https://doi.org/10.32614/CRAN.package.prismatic
Iannone, R., Cheng, J., Schloerke, B., Haughton, S., Hughes, E., Lauer, A., François, R., Seo, J., Brevoort, K., & Roy, O. (2026). *gt: Easily Create Presentation-Ready Display Tables*. https://doi.org/10.32614/CRAN.package.gt
Kassambara, A. (2026). *ggpubr: `ggplot2' Based Publication Ready Plots*. https://doi.org/10.32614/CRAN.package.ggpubr
Kassambara, A., Kosinski, M., & Biecek, P. (2026). *survminer: Drawing Survival Curves using `ggplot2'*. https://doi.org/10.32614/CRAN.package.survminer
Larmarange, J. (2025). *labelled: Manipulating Labelled Data*. https://doi.org/10.32614/CRAN.package.labelled
Larmarange, J., & Sjoberg, D. D. (2025). *broom.helpers: Helpers for Model Coefficients Tibbles*. https://doi.org/10.32614/CRAN.package.broom.helpers

Long, J. A. (2024). *interactions: Comprehensive, User-Friendly Toolkit for Probing Interactions*. https://doi.org/10.32614/CRAN.package.interactions

Lüdecke, D. (2018). sjmisc: Data and Variable Transformation Functions. *Journal of Open Source Software*, *3*(26), 754. https://doi.org/10.21105/joss.00754

Lüdecke, D. (2022). *sjlabelled: Labelled Data Utility Functions (Version 1.2.0)*. https://doi.org/10.5281/zenodo.1249215

Lüdecke, D., Ben-Shachar, M. S., Patil, I., & Makowski, D. (2020). Extracting, Computing and Exploring the Parameters of Statistical Models using R. *Journal of Open Source Software*, *5*(53), 2445. https://doi.org/10.21105/joss.02445

Lüdecke, D., Ben-Shachar, M. S., Patil, I., Waggoner, P., & Makowski, D. (2021). performance: An R Package for Assessment, Comparison and Testing of Statistical Models. *Journal of Open Source Software*, *6*(60), 3139. https://doi.org/10.21105/joss.03139

Lüdecke, D., Ben-Shachar, M. S., Patil, I., Wiernik, B. M., Bacher, E., Thériault, R., & Makowski, D. (2022). easystats: Framework for Easy Statistical Modeling, Visualization, and Reporting. *CRAN*. https://doi.org/10.32614/CRAN.package.easystats

Lüdecke, D., Patil, I., Ben-Shachar, M. S., Wiernik, B. M., Waggoner, P., & Makowski, D. (2021). see: An R Package for Visualizing Statistical Models. *Journal of Open Source Software*, *6*(64), 3393. https://doi.org/10.21105/joss.03393

Makowski, D., Lüdecke, D., Patil, I., Thériault, R., Ben-Shachar, M. S., & Wiernik, B. M. (2023). Automated Results Reporting as a Practical Tool to Improve Reproducibility and Methodological Best Practices Adoption. *CRAN*. https://doi.org/10.32614/CRAN.package.report

Mock, T. (2026). *gtExtras: Extending `gt' for Beautiful HTML Tables*. https://doi.org/10.32614/CRAN.package.gtExtras

Müller, K. (2025). *here: A Simpler Way to Find Your Files*. https://doi.org/10.32614/CRAN.package.here

Oberman, H. (2025). *ggmice: Visualizations for `mice' with `ggplot2'*. https://doi.org/10.32614/CRAN.package.ggmice

Ooms, J. (2025). *writexl: Export Data Frames to Excel `xlsx' Format*. https://doi.org/10.32614/CRAN.package.writexl

Ooms, J. (2026). *magick: Advanced Graphics and Image-Processing in R*. https://doi.org/10.32614/CRAN.package.magick

Patil, I. (2021). Visualizations with statistical details: The `ggstatsplot' approach. *Journal of Open Source Software*, *6*(61), 3167. https://doi.org/10.21105/joss.03167

Pedersen, T. L. (2025). *patchwork: The Composer of Plots*. https://doi.org/10.32614/CRAN.package.patchwork

Pedersen, T. L., Ooms, J., & Govett, D. (2026). *systemfonts: System Native Font Finding*. https://doi.org/10.32614/CRAN.package.systemfonts

Pedersen, T. L., & Shemanarev, M. (2026). *ragg: Graphic Devices Based on AGG*. https://doi.org/10.32614/CRAN.package.ragg

Ren, K., & Russell, K. (2021). *formattable: Create `Formattable' Data Structures*. https://doi.org/10.32614/CRAN.package.formattable

Robin, X., Turck, N., Hainard, A., Tiberti, N., Lisacek, F., Sanchez, J.-C., & Müller, M. (2011). pROC: an open-source package for R and S+ to analyze and compare ROC curves. *BMC Bioinformatics*, *12*, 77.

Robinson, D., Hayes, A., Couch, S., & Hvitfeldt, E. (2026). *broom: Convert Statistical Objects into Tidy Tibbles*. https://doi.org/10.32614/CRAN.package.broom

Rodriguez-Sanchez, F., & Jackson, C. P. (2025). *grateful: Facilitate citation of R packages*. https://pakillo.github.io/grateful/
Shi, B., Wang, Z., Choirat, C., & Valeri, L. (2026). *CMAverse: Causal Mediation Analysis*. https://github.com/BS1125/CMAverse
Sjoberg, D. D., Whiting, K., Curry, M., Lavery, J. A., & Larmarange, J. (2021). Reproducible Summary Tables with the gtsummary Package. *The R Journal*, *13*(1), 570–580. https://doi.org/10.32614/RJ-2021-053
Slowikowski, K. (2026). *ggrepel: Automatically Position Non-Overlapping Text Labels with `ggplot2'*. https://doi.org/10.32614/CRAN.package.ggrepel
Terry M. Therneau & Patricia M. Grambsch. (2000). *Modeling Survival Data: Extending the Cox Model*. Springer.
Therneau, T. M. (2026). *A Package for Survival Analysis in R*. https://CRAN.R-project.org/package=survival
Ushey, K., & Wickham, H. (2026). *renv: Project Environments*. https://doi.org/10.32614/CRAN.package.renv
Waring, E., Quinn, M., McNamara, A., Rubia, E. A. de la, Zhu, H., & Ellis, S. (2026). *skimr: Compact and Flexible Summaries of Data*. https://doi.org/10.32614/CRAN.package.skimr
Wickham, H. (2016). *ggplot2: Elegant Graphics for Data Analysis*. Springer-Verlag New York. https://ggplot2.tidyverse.org
Wickham, H. (2025a). *forcats: Tools for Working with Categorical Variables (Factors)*. https://doi.org/10.32614/CRAN.package.forcats
Wickham, H. (2025b). *stringr: Simple, Consistent Wrappers for Common String Operations*. https://doi.org/10.32614/CRAN.package.stringr
Wickham, H., & Bryan, J. (2026). *readxl: Read Excel Files*. https://doi.org/10.32614/CRAN.package.readxl
Wickham, H., Chang, W., Flight, R., Müller, K., & Hester, J. (2026). *sessioninfo: R Session Information*. https://doi.org/10.32614/CRAN.package.sessioninfo
Wickham, H., François, R., Henry, L., Müller, K., & Vaughan, D. (2026). *dplyr: A Grammar of Data Manipulation*. https://doi.org/10.32614/CRAN.package.dplyr
Wickham, H., & Henry, L. (2026). *purrr: Functional Programming Tools*. https://doi.org/10.32614/CRAN.package.purrr
Wickham, H., Henry, L., Pedersen, T. L., Luciani, T. J., Decorde, M., & Lise, V. (2025). *svglite: An `SVG' Graphics Device*. https://doi.org/10.32614/CRAN.package.svglite
Wickham, H., Pedersen, T. L., & Seidel, D. (2025). *scales: Scale Functions for Visualization*. https://doi.org/10.32614/CRAN.package.scales
Wilke, C. O. (2025). *cowplot: Streamlined Plot Theme and Plot Annotations for `ggplot2'*. https://doi.org/10.32614/CRAN.package.cowplot
Wilke, C. O., & Wiernik, B. M. (2022). *ggtext: Improved Text Rendering Support for `ggplot2'*. https://doi.org/10.32614/CRAN.package.ggtext
Wood, S. N. (2003). Thin-plate regression splines. *Journal of the Royal Statistical Society (B)*, *65*(1), 95–114. https://doi.org/10.1111/1467-9868.00374
Wood, S. N. (2004). Stable and efficient multiple smoothing parameter estimation for generalized additive models. *Journal of the American Statistical Association*, *99*(467), 673–686. https://doi.org/10.1198/016214504000000980
Wood, S. N. (2011). Fast stable restricted maximum likelihood and marginal likelihood estimation of semiparametric generalized linear models. *Journal of the Royal Statistical Society (B)*, *73*(1), 3–36. https://doi.org/10.1111/j.1467-9868.2010.00749.x

Wood, S. N. (2017). *Generalized Additive Models: An Introduction with R* (2nd edn). Chapman and Hall/CRC.
Wood, S. N., Pya, N., & Säfken, B. (2016). Smoothing parameter and model selection for general smooth models (with discussion). *Journal of the American Statistical Association*, *111*, 1548–1575. https://doi.org/10.1080/01621459.2016.1180986
Xie, Y. (2014). knitr: A Comprehensive Tool for Reproducible Research in R. In V. Stodden, F. Leisch, & R. D. Peng (Eds), *Implementing Reproducible Computational Research*. Chapman and Hall/CRC.
Xie, Y. (2015). *Dynamic Documents with R and knitr* (2nd edn). Chapman and Hall/CRC. https://yihui.org/knitr/
Xie, Y. (2025). *knitr: A General-Purpose Package for Dynamic Report Generation in R*. https://yihui.org/knitr/
Xie, Y., Allaire, J. J., & Grolemund, G. (2018). *R Markdown: The Definitive Guide*. Chapman and Hall/CRC. https://yihui.org/rmarkdown/
Xie, Y., Dervieux, C., & Riederer, E. (2020). *R Markdown Cookbook*. Chapman and Hall/CRC. https://yihui.org/rmarkdown-cookbook
